# AI-Generated Responses to Patient’s Messages: Effectiveness, Feasibility and Implementation

**DOI:** 10.64898/2026.03.02.26347175

**Authors:** Kim J.M. Bladder, Arie C. Verburg, Milou Arts-Tenhagen, René Willemsen, Guido B. van den Broek, Chantal M.L. Driessen, Rieke J.B. Driessen, Bas Robberts, Arthur R.T. Scheffer, Arjen P. de Vries, Tim Frenzel, Julie E.M. Swillens

## Abstract

**Background:** Generative artificial intelligence (GenAI) in healthcare may reduce administrative burden and enhance quality of care. Large language models (LLMs) can generate draft responses to patient messages using electronic health record (EHR) data. This could mitigate increased workload related to high message volumes. While effectiveness and feasibility of these GenAI tools have been studied in the United States, evidence from non-English contexts is scarce, particularly regarding user experience.

**Objective:** This study evaluated the effectiveness, feasibility and barriers and facilitators of implementing Epic’s Augmented Response Technology (Art) GenAI tool (Epic Systems Corporation, Verona, WI, USA) in a Dutch academic healthcare setting among a broad range of end users. It explored healthcare professionals’ (HCP) usage metrics, expectations, and early user experiences.

**Methods:** We conducted a hybrid type 1 effectiveness-implementation design. HCPs of four clinical departments (dermatology, medical oncology, otorhinolaryngology, and pulmonology) participated in a six-month study. Effectiveness of Art was assessed using efficiency indicators from Epic (including all InBasket users in the hospital) and survey scales measuring well-being and clinical efficiency at three time points: PRE, POST-1 (1 month), and POST-2 (4 months). Feasibility of Art was evaluated through adoption indicators from Epic and survey scales on use and usability. Barriers and facilitators of Art implementation were collected through the survey and thematized using the NASSS framework (Nonadoption, Abandonment, Scale-up, Spread and Sustainability).

**Results:** 237 unique HCPs generated a total of 8,410 drafts. Review and drafting times were similar for users with and without Art, indicating minimal differences. Perceived clinical efficiency declined significantly from PRE to POST-2, while well-being remained unchanged. Adoption was initially high but decreased over time, averaging 16.7% across departments. Usability and intention-to-use scores also declined significantly. Qualitative findings highlighted time savings, well-structured drafts, and patient-centered language as facilitators. Reported barriers included limited impact on time, low practical utility, content inaccuracies, and style misalignment.

**Conclusions:** This evaluation of a GenAI tool for patient-provider communication in a non-English academic hospital revealed mixed perceptions of effectiveness and feasibility. High initial expectations contrasted with limited perceived impact on time-savings, well-being and clinical efficiency, alongside declining adoption and usability. Barriers and facilitators revealed contrasting views. These findings underscore the need for a workflow for the handling of user feedback, guidance on clinical responsibilities, along with clear communication about the tool’s purpose and limitations to manage expectations. Additionally, establishing consensus on a set of quality indicators and their thresholds that indicate when a GenAI tool is sufficiently robust will be critical for responsible scaling of GenAI in clinical practice.

## Introduction

Generative artificial intelligence (GenAI) is rapidly gaining momentum in healthcare. These technologies generate new content based on patterns learned from existing data and hold substantial promise to enhance quality of care and reduce workload for healthcare professionals (HCPs) [1–4]. GenAI is being explored for a range of applications, including clinical documentation assistance, providing patient information, and supporting billing processes [3].

GenAI shows promise in reducing the growing administrative burden on HCPs [5]. One contributor to the burden is the increasing volume of patient-provider messages. During the COVID-19 pandemic, the shift toward telemedicine in patient-provider care resulted in a 157% increase in patient messages [6]. This increased volume sustained after the pandemic [7]. Previous studies have found a correlation between high message volume and increased risks of burnout and exhaustion, showing the need to reduce message-related workload [8,9].

Large language models (LLMs) offer a promising solution to support HCPs in reducing the workload associated with high message volume by generating draft responses to patient messages using contextual Electronic Health Record (EHR) data [10]. By leveraging relevant clinical information from the EHR to create a draft reply, these models could reduce cognitive load and help improve the usability of patient-provider communication systems [11]. This, in turn, can help relieve the administrative burden.

Despite the potential of LLMs, their real-world clinical impact remains largely unexamined [12]. While adoption and user experiences with these tools have been explored in the United States [10,13–15], little is known about how such tools are received in non-English healthcare settings. A recent study from the University Medical Center Groningen demonstrated that a safe implementation of LLM-generated draft responses in a non-English healthcare setting using Epic’s Augmented Response Technology (Art, Epic Systems Corporation, Verona, WI, USA) is viable [16]. However, this study did not yet examine user experiences specifically, so it remains unclear how non-English HCPs perceive such tools in daily clinical practice.

To address this gap, this study aimed to assess the effectiveness and feasibility of Art, as well as barriers and facilitators to its implementation in clinical practice, by exploring usage metrics, expectations, and early user experiences.

## Methods

We used a hybrid type 1 design to evaluate effectiveness, feasibility, and barriers and facilitators of Art simultaneously [17]. We collected usage metrics via the EHR and conducted a repeated measures survey study, to assess expectations and early user experiences with Art. This study was reported in accordance with the CHERRIES (Checklist for Reporting Results of Internet E-Surveys) [18]. A complete CHERRIES checklist is provided in Multimedia Appendix 1.

### Integration of Art

At Radboud University Medical Center (Radboudumc), the GenAI tool Art was enabled into the EHR in October 2024. This tool supports HCPs by deploying an LLM to generate a draft reply to a patient message, within the patient-provider communication system InBasket. Figure S1 in Multimedia Appendix 2 displays a visualization of the Art user interface and Figure S2 illustrates a schematic of the technological workflow of Art.

Art performed the following workflow to produce its draft responses, also referred to as ‘drafts’ throughout this study. Every incoming patient message of type 54 (medical question from patient to HCP) was classified into 4 categories using GPT-3.5 Turbo [19]: administrative, general, medication, and results. Each category was linked to a specific prompt that determined which contextual information should be retrieved from the EHR to generate a draft. These prompts were initially provided by Epic Systems Corporation and were subsequently refined based on insights from frontrunner hospitals in Art within the Netherlands. An IT Application Specialist (RW) in consultation with a Medical Information Officer (TF) performed additional limited prompt engineering, incorporating feedback from the 23 HCPs who tested the tool during the initial validation phase. These refinements included ensuring consistent closing, adding the name of the HCP and aligning with the usual communication style of Radboudumc. At the end of February 2025, two additional adjustments were implemented based on user feedback: automatically adding the provider’s name to the closing and extending the lab values retrieval window in the result category prompt from 14 to 60 days.

The draft message itself was automatically generated using GPT-4o [20], which operated as a static model and did not learn from any input data. If any issue occurred during message processing (e.g., different language, attachments, or message changes), no draft would appear for the user. HCPs using InBasket could choose to initiate their response with the LLM-generated draft, or discard the draft and compose a reply from scratch. Additionally, Art users had the option to provide feedback on each generated draft. Using buttons and text, each user could identify a draft as useful and give constructive feedback in 5 categories: factual correctness, relevance, quality of patient guidance, incorrect name receiver, and length (too long or too short).

### Study Design

We extracted and analyzed InBasket data from the EHR and simultaneously conducted a survey-based evaluation, to determine the effectiveness, feasibility, and implementation of Art. Usage metrics from the EHR were collected during the study period from January to June 2025. The survey-based evaluation consisted of a baseline (pre-implementation) and two follow-up (post-implementation) measurements at one and four months after Art implementation. The study timeline is displayed in Figure 1. The core project team consisted of a professor in data science (AdV), a junior AI researcher (KB), and three senior researchers: one with a clinical focus (TF), one with an implementation research focus (JS), and one with a focus on clinimetrics (AV).

**Figure 1.**
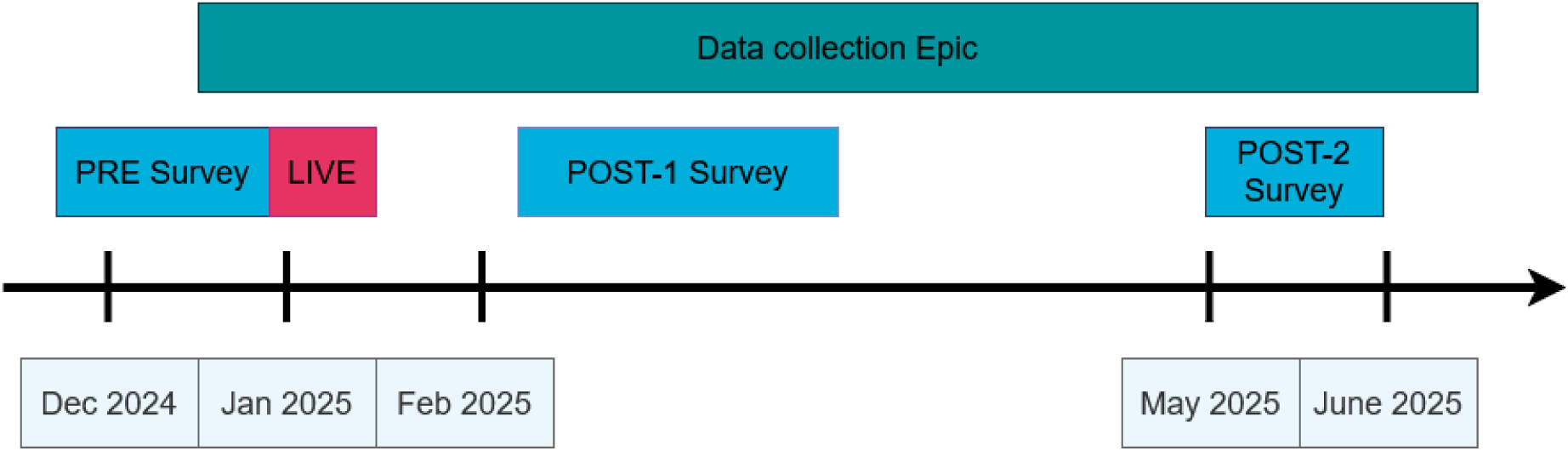
Timeline of the measurements and study period.

### Study Population and Data Collection

The study involved four clinical departments of Radboudumc: dermatology, pulmonology, medical oncology, and otorhinolaryngology (ENT). HCPs were introduced to Art by email and oral communication, provided by the Medical Information Officer (MIO) associated with the department as ambassador for the project. A tip sheet was provided to guide users on how to use Art and how to submit feedback on generated drafts. During the study, the ambassadors and heads of departments were kept informed about the usage metrics of the tool and were asked to remind their colleagues to make use of Art, complete the survey, and provide feedback.

Data on effectiveness and feasibility were collected from both the EHR and the survey. Data on barriers and facilitators were only collected by the survey.

#### Study Population and Data Collection from EHR

The EHR vendor configured predefined usage metrics including efficiency indicators and adoption indicators. Data was aggregated at department level, and no data at the individual user or patient level was included in the dataset. The efficiency indicators included all messages from all Radboudumc InBasket users (n=11,126) between January and June 2025. The adoption indicators were collected among all users of Art (n=237) during the study period.

#### Study Population and Data Collection from Survey

All types of HCPs that work with InBasket at the four departments were eligible for the study and were divided into three categories: physicians, nurses, and support staff. A total of 148 HCPs were eligible for participation in the PRE-measurement, 164 for POST-1, and 158 for POST-2. Fluctuations in participant numbers were caused by professionals being out of office for a longer period or no longer employed at the time of data collection.

Closed e-surveys were developed using LimeSurvey (version 6.5.4) and technical validation of survey was checked by the project team. The surveys were distributed by email together with an information letter that outlined the study’s purpose, identified the researchers, specified the estimated completion time (10-15 minutes), and explained how participants’ data would be handled. Each participant received a unique link in the invitation email, which allowed them to access and complete the survey. They could also return later to finish the survey using the same link and were able to navigate back to previous questions and modify their responses until the survey was closed. Completeness of a survey was checked by highlighting mandatory items in LimeSurvey. Initially, participants were only given access to Art after completing the pre-implementation survey. However, all HCPs across the participating departments were eventually granted access to the tool at the end of January 2025 (n=237). Participation in the surveys was voluntary, and no additional incentives were offered.

### Study Measures

#### Measures from EHR

The effectiveness of the tool was measured using efficiency indicators, specifically review time and draft time. Review time was defined as the time between opening a patient’s message and starting a reply, while draft time referred to the time spent composing a reply after initiating the draft with either a blank reply or the generated draft. For these efficiency indicators, a distinction was made between messages that used Art and messages where no draft was generated by Art. The latter included messages from users from other departments where Art was not enabled or messages where no draft was generated.

Additionally, feasibility was assessed using data from the EHR on adoption indicators. These included number of generated drafts, adoption rate, and feedback counts. A draft was considered used when the user chose to initiate their response with it, regardless of how the content was subsequently modified. The number of feedback items per category was gathered and analyzed to gain insight into specific areas for improvement. Table S1 in Multimedia Appendix 2 summarizes all metrics.

#### Measures from Survey

Based on insights drawn from literature on theories, models and frameworks for evaluating information technology in healthcare and available expertise within the project team, a survey was composed to assess the expectations (PRE survey) and experiences (POST survey) on the effectiveness, feasibility and implementation of Art. Table S2 in Multimedia Appendix 2 presents an outline of the survey. The first part of the survey consisted of questions gathering the demographic characteristics of the participants, including general trust in AI [21–23]. The remaining sections of the survey covered four domains, two to assess effectiveness: Well-being and Clinical Efficiency, and two to assess feasibility: Use of Art and Usability of Art [12,13,24–26].

For effectiveness, Well-being was evaluated using the NASA Task Load Index [27] and the work exhaustion subscale of the Professional Fulfillment Index (PFI-WE) [28]. We used the Usefulness scale from the Technology Acceptance Model (TAM-PU) [25] to measure the impact of Art on Clinical Efficiency. Motivated by previous research on Art effectiveness, we supplemented the domain with a custom item on expected and perceived effects [13]. An open-ended question was included to further explore the added value of Art on Clinical Efficiency.

Feasibility was assessed through the domain Use of Art, which included questions to indicate how frequently participants used the tool after implementation, both in the past week and in total. Art use was defined as subjectively having used Art at least once since given access. Participants who reported no use were asked to provide a short explanation to help understand why they had not used Art. Behavioral intention to use was assessed with the TAM-BI scale [25] and Attitude towards Art (TAM-ATT) [25] was measured with a separate scale in the PRE survey. The domain Usability of Art also measured feasibility by including the ease-of-use scale (TAM-PEOU) [25]. Additionally, we added a custom item based on previous research to collect the expected and perceived quality of the generated output [26]. The System Usability Scale (SUS) [29] and the Net Promotor Score (NPS) [30] assessed user satisfaction in the POST-1 and POST-2 measurements. SUS scores range from 0 to 100. Scores above 68 are generally considered above average, with higher scores indicating better usability. NPS ranges from −100 to +100. Negative values indicate more detractors (respondents who would not recommend the tool) than promoters (those who would). Positive scores are considered good, with scores above 50 often interpreted as excellent. All outcomes, except for NASA Task Load Index and NPS scale, were rated on a 5-point Likert scale (1 = totally disagree to 5 = totally agree).

The survey concluded with open-ended questions to explore barriers and facilitators to Art implementation, and to receive suggestions for improvement. These questions were included as an essential step in early implementation research [17]. They were open-ended because the scientific literature at that time did not yet provide information with respect to barriers and facilitators of GenAI in clinical practice. A final open-ended question served the goal to gather future perspectives on the use of fully automatic response systems. In total, the survey consisted of six pages: one page for informed consent, one page for participant demographics (11 items), two pages containing the four domain scales and open-ended questions with a total of 17 items distributed across them, one page for future perspectives (1 item), and a final closing page.

To assess the internal consistency and structural validity of the survey items, Cronbach’s alpha coefficients were calculated, and confirmatory factor analyses (CFA) were conducted for each scale. Table S3 in Multimedia Appendix 2 displays the Cronbach’s alpha coefficients across the three measurements. Confirmatory factor analyses (CFA) supported the structural validity of most scales as shown in Table S4 in Multimedia Appendix 2.

### Analysis

All analyses were performed using Python software (version 3.13.5; Python Software Foundation).

#### EHR analysis

Usage metrics extracted from Epic (efficiency indicators and adoption indicators) were summarized using descriptive statistics. Underlying data and measures of variability were not available, rendering statistical significance testing impossible.

#### Survey analysis Quantitative Analysis

We captured survey responses automatically in LimeSurvey and only analyzed completed questionnaires. For each outcome scale, descriptive statistics (mean and standard deviation) were calculated per timepoint. To ensure an accurate representation in the subsequent analyses, we only included participants who filled out the survey but did not use Art in the analysis of the barriers and facilitators of the POST measurements.

We analyzed changes over time (PRE, POST-1, POST-2) using linear mixed model analysis, with a random intercept for respondents and a fixed effect for time. No random slopes were included, and the residual covariance structure was assumed to be independent (default in the Python package `statsmodels`; version 0.14.4). For comparisons between two timepoints, ordinary least squares (OLS) regression was used instead, using the Huber-White standard errors. All available responses per timepoint were included in the analyses, as comparison of participant demographic characters across timepoints did not show selective dropout.

The Shapiro-Wilk test assessed the normality assumption of model residuals. Given that this test is highly sensitive to minor deviations, additional visual inspections were performed using QQ-plots. For scales that suggested non-normality, additional regression analyses were conducted using robust Huber-White standard errors. The results only showed minimal differences in effect estimates. Considering that linear mixed models are generally robust to mild violations, and that robust analyses did not lead to different conclusions, the original model results were reported.

Due to limited statistical power within subgroups, differences between departments and professions were not included in the mixed model analysis and are only presented descriptively.

#### Qualitative Analysis

Answers to open-ended questions from POST-1 and POST-2 were aggregated for analysis, whereas PRE responses were analyzed separately. These answers were then coded, starting with framework coding and supplemented with thematic coding. We used the NASSS framework (Nonadoption, Abandonment, Scale-up, Spread and Sustainability) to categorize the barriers and facilitators into the domains (illness/condition, technology, value proposition, adopter system, organization(s), wider context, and embedding and adaptation over time) [31]. Initial coding was performed by the primary researcher (KB), after which the codes and categorization were reviewed and refined in collaboration with a second researcher (AV). Consensus was reached through discussion to ensure consistency and reliability in theme interpretation. In case of a discrepancy, a third researcher (JS) was consulted.

### Ethical Considerations

Data exchange between the EHR system and the LLM hosting environment takes place under strict contractual agreements between the hospital and the EHR vendor, complying with privacy regulations. Submitted data is never used for model training, and human review by external parties is not permitted.

This study did not require approval of an institutional review board under Dutch National Law (Medical Research Involving Human Subjects Act (WMO)). The patient advisory council was consulted prior to implementation, and they expressed support for evaluating GenAI initiatives. The study participants gave informed consent prior to participation in the survey. Personal contact information used for communication was stored separately from research data. Research data was pseudonymized before analysis.

## Results

### Usage metrics: Data from EHR

Efficiency indicators were derived from all active Radboudumc InBasket users (n = 11,126). Demographic data was not available for the efficiency indicators. Adoption indicators were based on Epic data covering patient-provider messages exchanged by 237 HCPs from four participating departments during the January–June 2025 study period. This data originated from 55 professionals from Dermatology, 51 from ENT, 59 from Pulmonology, and 72 from Medical Oncology.

Descriptive statistics showed that the average review time for patient messages with access to Art was slightly different than for messages without access (2 minutes and 2 seconds and 2 minutes and 10 seconds, respectively). Similarly, draft time showed a small change for messages written with Art (1 minute and 52 seconds versus 1 minute and 56 seconds).

Adoption indicators demonstrated that Art generated a total of 8,410 drafts during the study period, of which 1,401 drafts (16.7%) were used across all departments. The pulmonology department used the most drafts (598 drafts), while the ENT department had the lowest number of drafts used (171 drafts). Pulmonology had the highest adoption rate (27.1%) and the medical oncology department the lowest (10.7%). The ENT department and dermatology department had 17.8% and 17.1% adoption rate, respectively. The department of pulmonology and medical oncology exhibited the largest decrease in adoption rate. The dermatology department displayed a small decrease over time, while the department of ENT was consistent in using Art. Figure 2 presents the percentual use of drafts per department. Table S5 in Multimedia Appendix 2 displays an extended overview of the total adoption of Art per department over 6 months.

**Figure 2.**
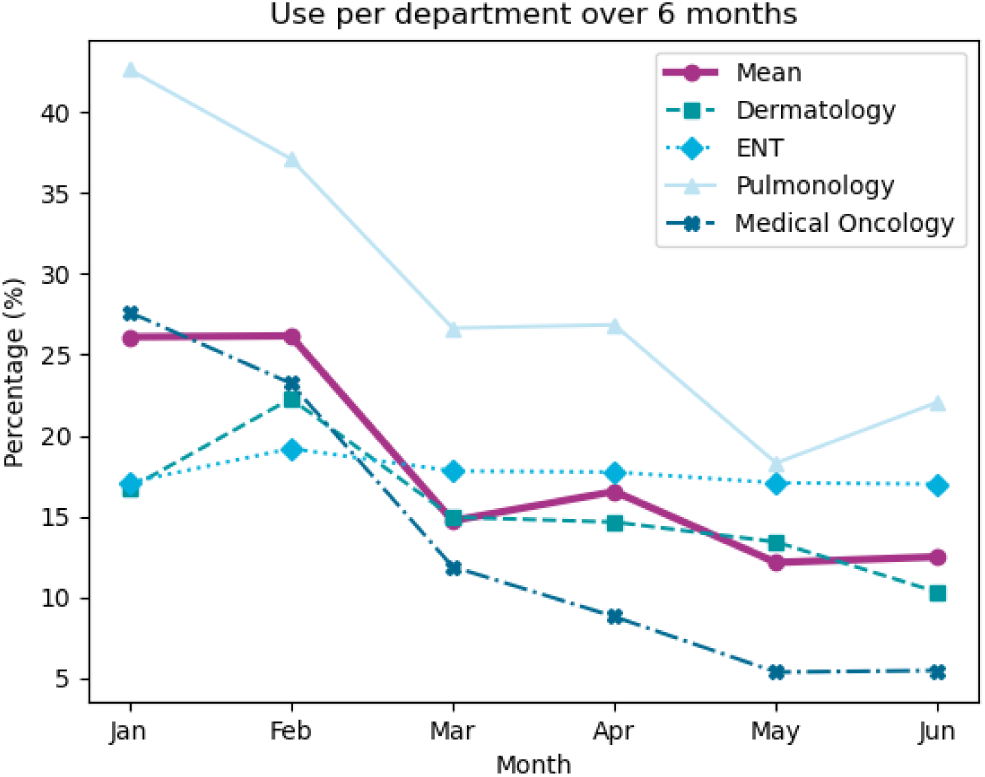
Percentual use of drafts per department over a 6-month period. The figure displays the percentage of drafts used per department, as well as the overall average adoption rate across departments.

**Figure 3.**
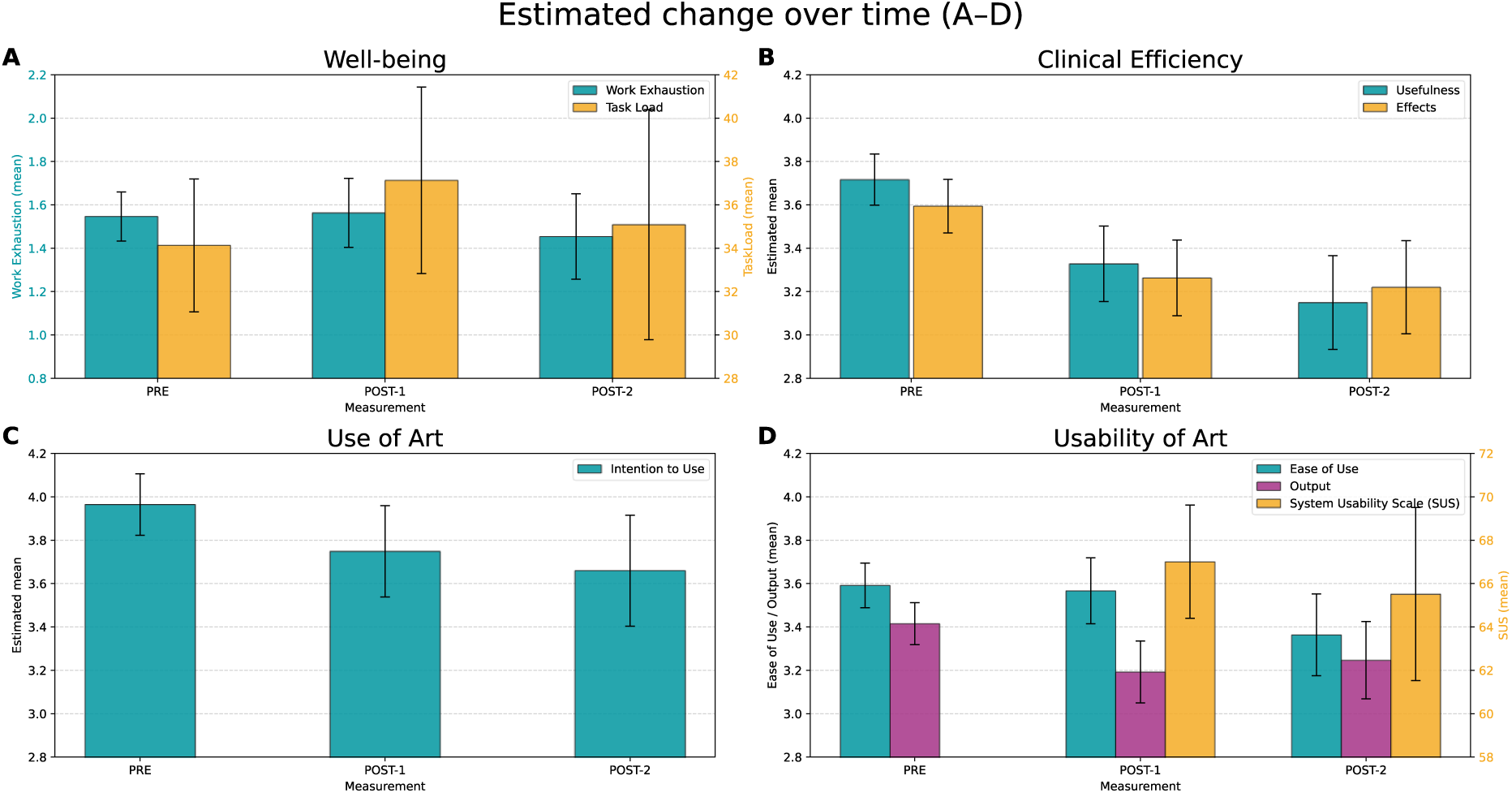
Estimated change over time of the domains (a) Well-being, (b) Clinical Efficiency, (c) Use of Art, and (d) Usability of Art. The figures display mean scores with S5% confidence intervals. PRE represents expectations, whereas POST (1 and 2) represent experienced scores. All scales are in range 1-5 except for the Task Load scale (figure a) and the System Usability Scale (figure d), which both range from 0-100.

In the 6 months of the study, 186 drafts (2.2%) were identified as useful, and for another 424 drafts (5.0%), constructive feedback was given. Of the latter, 37.5% concerned factual incorrectness, 30.4% addressed lack of relevance, 5.6% related to quality of patient guidance, 5.0% to incorrect recipient name, and 4.2% to length. Figure S2 in Multimedia Appendix 2 shows the distribution over 6 months and per department for both positive and constructive feedback.

### Survey

#### Participant Demographics

From the included HCPs, 108 participants (73.0%) completed the PRE-survey, 58 (35.4%) completed the POST-1 survey, and 37 (23.4%) completed the POST-2 survey. Art use, defined as having used Art at least once, was reported by 46 participants (79.3%) at POST-1 and 29 (78.4%) at POST-2; only these participants were included in the mixed model analysis of the surveys.

Reasons for not using Art included a lack of time to explore the tool or not having to respond to digital patient messages in the past period. Other participants indicated that they had reviewed the drafts but chose not to use the draft, citing issues such as insufficient comprehension of the patient’s questions by Art, incomplete drafts, digital stress, or a tendency to revert to familiar routines. Table 1 presents the demographic characteristics of the study population used for qualitative analysis.

**Table 1.** Characteristics of the study population for each time point. Only participants that reported to have used Art at least once were included in these characteristics.

|  | PRE (n=108) | POST-1 (n=46) | POST-2 (n=29) |
| --- | --- | --- | --- |
| **Gender** |  |  |  |
| Female | 88 (81.5%) | 39 (84.8%) | 22 (75.9%) |
| Male | 20 (18.5%) | 7 (15.2%) | 7 (24.1%) |
| Age<sup>+</sup> | 44.8 (11.4) | 43.4 (11.3) | 45.9 (10.2) |
| Work experience<sup>**</sup> (profession) | 12.1 (10.9) | 12.8 (11.1) | 12.9 (10.2) |
| Work experience<sup>**</sup> (total) | 19.3 (11.8) | 19.0 (11.1) | 19.9 (11.4) |
| Experience with Epic<sup>**</sup> | 8.5 (3.6) | 8.6 (4.1) | 8.9 (3.2) |
| Experience with In Basket<sup>**</sup> | 5.9 (3.5) | 6.6 (3.5) | 7.4 (3.5) |
| Interest in GenAI within work environment<sup>**</sup> | 4.1 (0.7) | 4.1 (0.5) | 4.0 (0.7) |
| Interest in GenAI outside work environment<sup>**</sup> | 3.6 (0.9) | 3.6 (0.9) | 3.4 (0.9) |
| Trust in AI<sup>**</sup> | 3.3 (0.5) | 3.3 (0.4) | 3.1 (0.5) |
| **Department** |  |  |  |
| Dermatology | 31 (28.7%) | 18 (39.1%) | 7 (24.1%) |
| Ear, Nose and Throat | 15 (13.9%) | 6 (13.0%) | 6 (20.7%) |
| Pulmonology | 19 (17.6%) | 11 (23.9%) | 11 (37.9%) |
| Medical Oncology | 43 (39.8%) | 11 (23.9%) | 5 (17.2%) |
| **Categorized type of profession** |  |  |  |
| Physician | 56 (51.9%) | 23 (50.0%) | 20 (69.0%) |
| Nurse | 40 (37.0%) | 16 (34.8%) | 9 (31.0%) |
| Support staff | 12 (11.1%) | 7 (12.1%) | 0 (0.0%) |

<sup>+</sup>mean (SD); <sup>\*</sup>in years; <sup>†</sup>items reflect responses on a 1-5 Likert scale.

#### User Expectations and Experiences

User expectations and experiences were assessed across four domains: Well-being, Clinical Efficiency, Use of Art, and Usability of Art. Figure 1 visualized the changes over time, showing the estimated coefficients and their 95% confidence intervals for each outcome scale. Tables S6-S9 in Multimedia Appendix 2 displays the descriptive statistics for all scales and subgroups. Table S10 in Multimedia Appendix 2 shows an extended overview of all changes over time.

#### Effectiveness

For the domain Well-being, consisting of task load and work exhaustion, analyses revealed no significant changes.

Significant declines over time were observed across both measures of Clinical Efficiency. Perceived usefulness of Art decreased between PRE and POST-1 (coefficient = –0.39; 95% CI, – 0.58 to –0.20; p<.001), and between PRE and POST-2 (coefficient = –0.57; 95% CI, –0.80 to – 0.34; p<.001). Perceived effects followed a similar downward pattern, with significant reductions between PRE and POST-1 (coefficient = –0.33; 95% CI, –0.51 to –0.15; p<.001), and PRE and POST-2 (coefficient = –0.37; 95% CI, –0.60 to –0.15; p<.001).

#### Feasibility

Mixed model analyses indicated a decrease in expected and perceived (POST-2) intention to use (coefficient = –0.30; 95% CI, –0.58 to –0.03; p=.03). No significant difference between PRE and POST-1 was found.

For Usability of Art, ease-of-use decreased from PRE to POST-2 (coefficient = –0.23; 95% CI, – 0.43 to –0.03; p=.03) but showed no significant decline between PRE and POST-1. In contrast, a significant decrease between PRE and POST-1 was recorded for perceived output (coefficient = –0.22; 95% CI, –0.38 to –0.07; p=.005), whereas the decline between PRE and POST-2 was not statistically significant. We found no significant change between POST-1 and POST-2 for the SUS scale. The NPS decreased from POST-1 to POST-2 (–13.04 to –37.93), indicating a decline in participants’ intention to recommend.

#### Barriers and Facilitators

The barriers, facilitators, and suggestions for improvement were clustered in themes. Table 2 provides an overview, together with representative quotations.

**Table 2.**
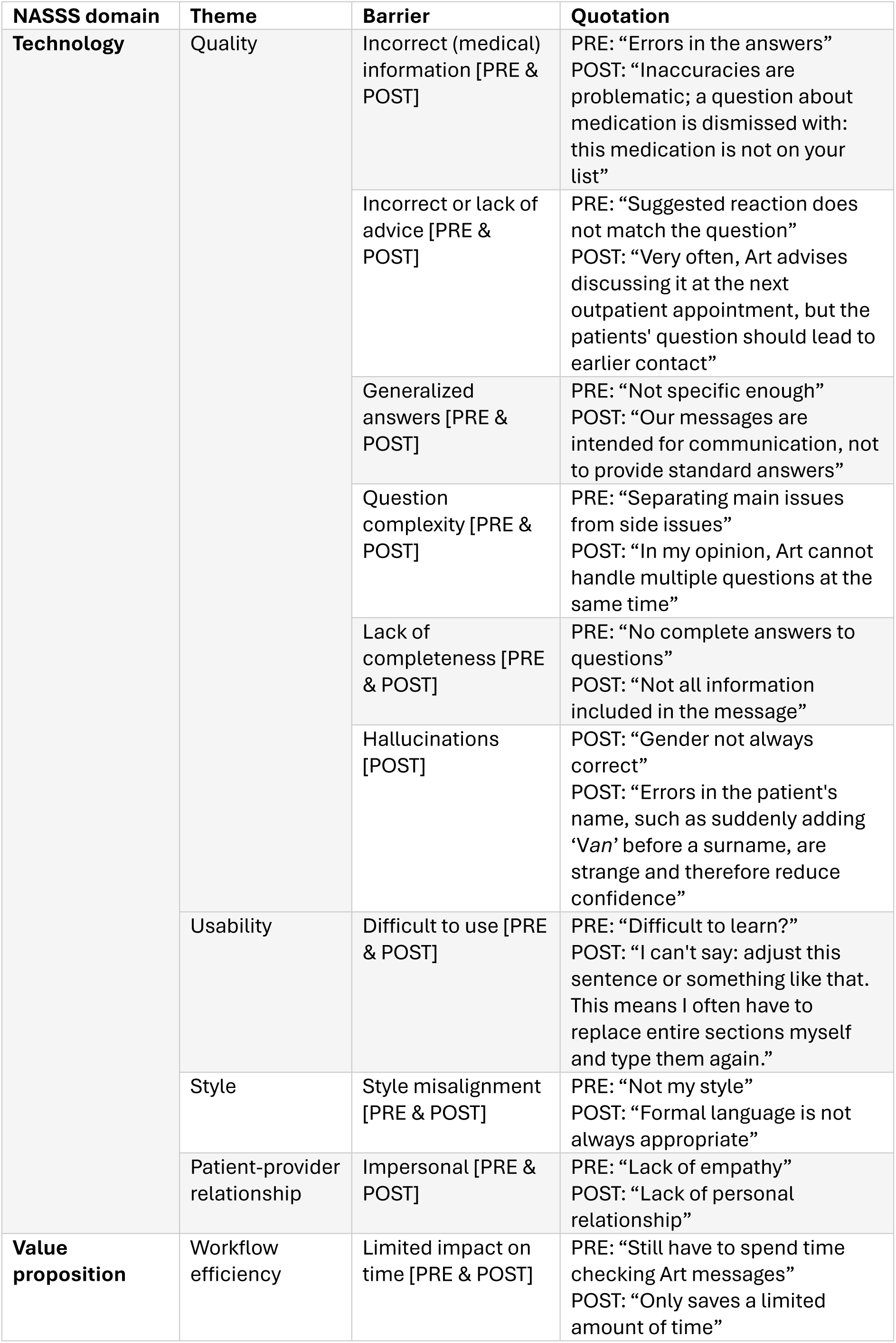
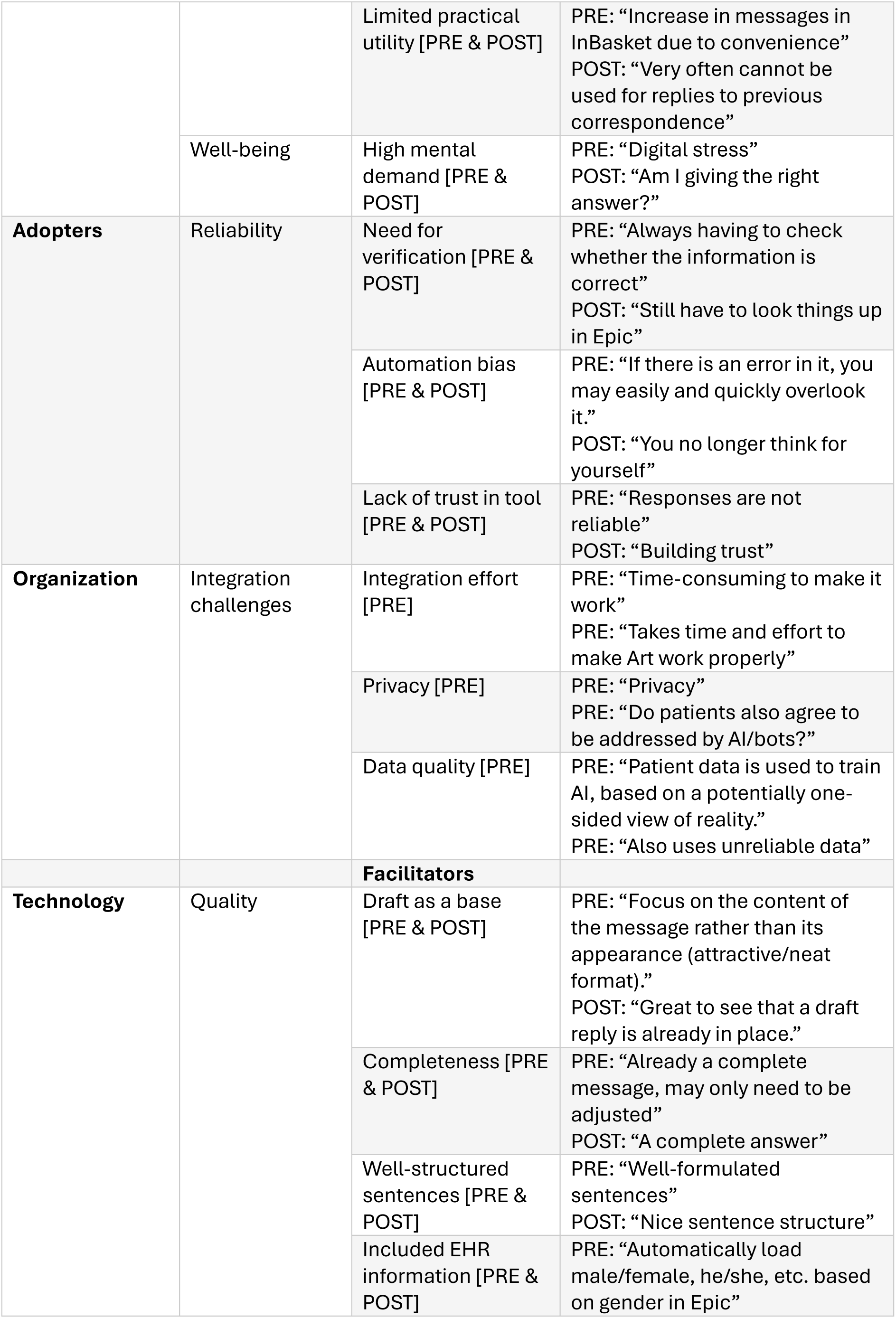
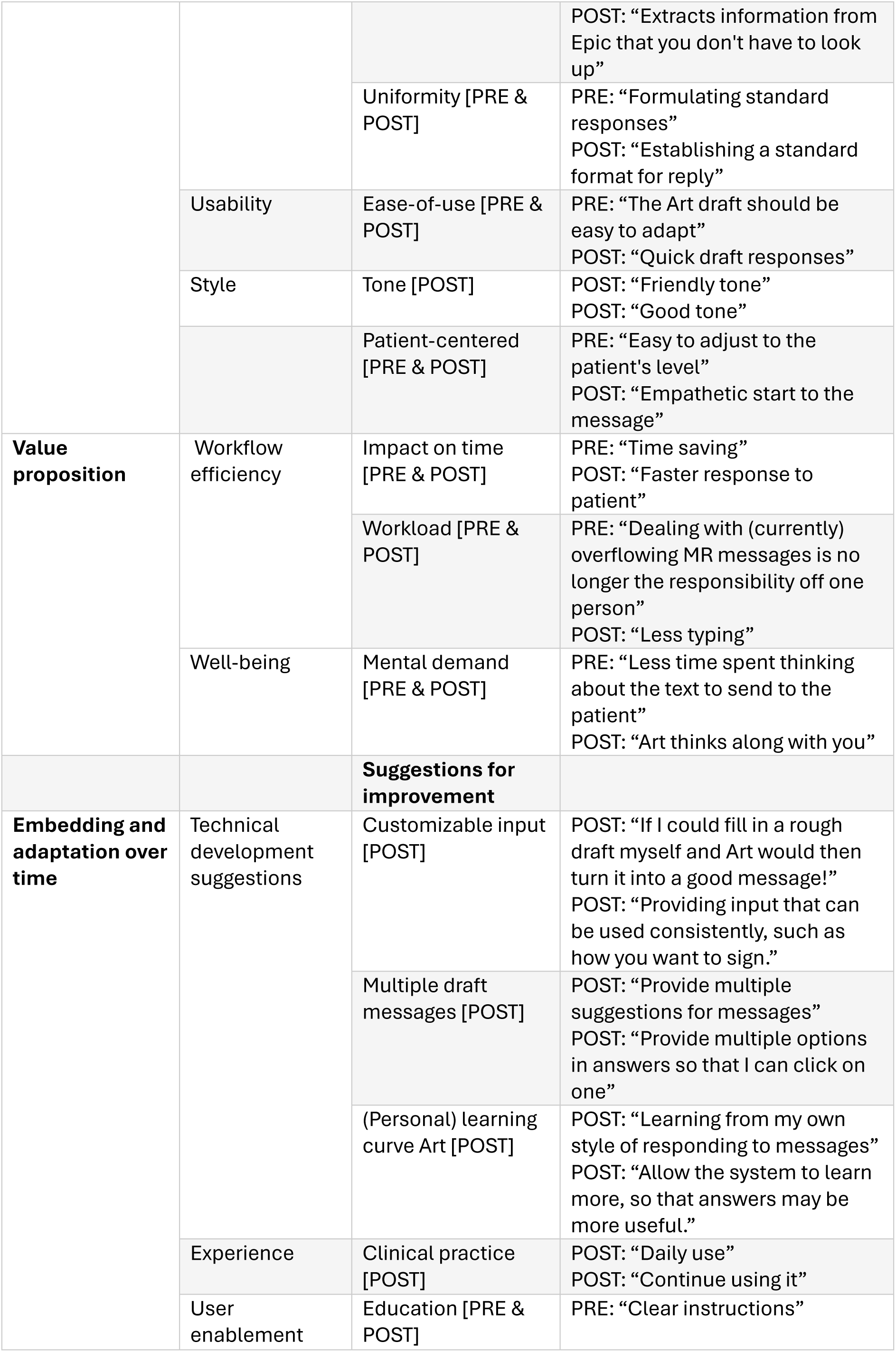
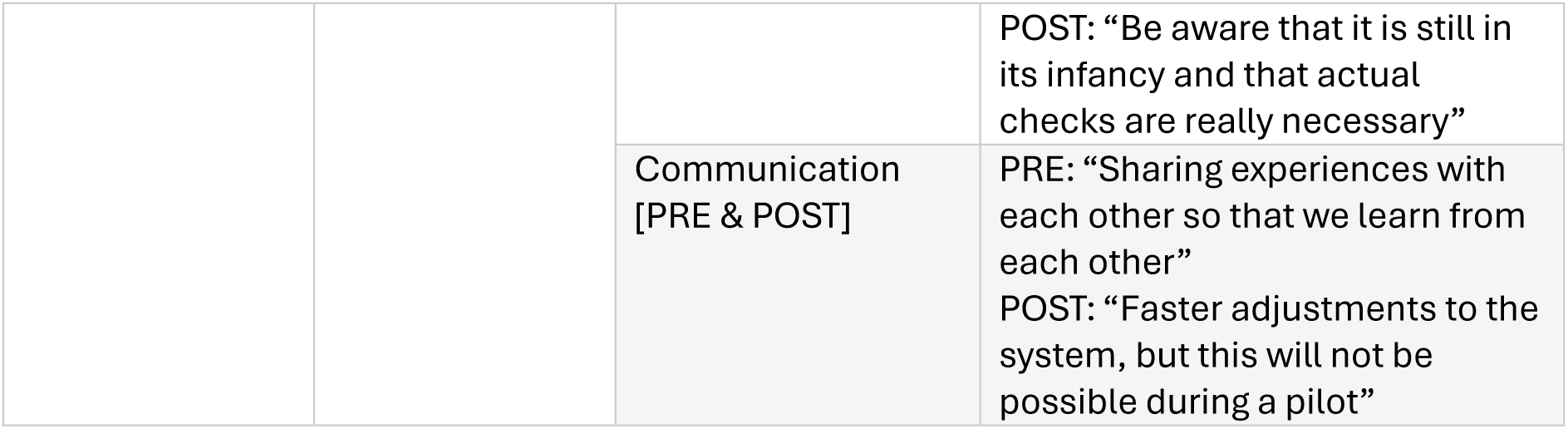
Barriers, Facilitators and suggestions for improvements. The quotes are translated from Dutch.

#### Barriers

Within the technology domain, barriers surrounded the quality and usability of the content, with people expecting errors in answers, incompleteness, and a steep learning curve for using the tool. Respondents indicated that they indeed experienced many medical inaccuracies in the draft messages. Users also perceived Art to be unable to handle multiple questions when asked in a single patient message. Additionally, participants expressed discomfort on the patient-provider relationship using Art given the inherently impersonal nature of such an application.

The value proposition is centered around the workflow efficiency and the negative effect on well-being of users when having to adapt to a new system. Users anticipated that the impact on time would be limited as they still need to check the contents of the draft. Experiences indeed highlighted limited time savings and limited practical utility. Regarding the adopter domain, barriers that were expected were also present after implementation of Art. These barriers were linked to reliability, as some people fear that mistakes by Art could easily be overlooked and worried about automation bias, where users might blindly trust what the LLM generates. Before implementation, concerns on privacy, integration effort, and data quality were raised regarding integration challenges in the organization domain. However, these were not expressed after Art implementation.

#### Facilitators

The technology domain in facilitators consisted mostly of the usability of the tool, and quality of the draft replies. Users expected that drafts would allow users to focus more on the content of the message, rather than the tone and the layout of messages. Their experiences also showed that the drafts were quick to use and provided a good basis for a response. Additionally, they anticipated that a draft would help the patient by being able to adjust to the patient’s level. After implementation, users indicated a sufficient and empathic style of the generated draft.

Regarding value proposition, common expectations before implementation for workflow efficiency included time savings and reducing the burden of managing overflowing message inboxes. After using the tool, most respondents mentioned efficiency benefits such as quicker responses to patients and reduced typing. Additionally, people indicated a positive influence on their well-being as they anticipated and perceived a reduction in mental workload.

#### Suggestions for improvements

All suggestions for improvement were related to the embedding and adaption over time domain. Suggestions for improvement in technology focused on personalization of input and output. Participants expressed a preference for a tool that could provide multiple draft suggestions or could adjust their own rough drafts into well-written messages, as well as a tool that could learn from their response styles and previous correspondence. Concerning adopters, the need was mentioned to gain more experience in clinical practice. Related to organization, users indicated the need for effective user enablement with clear instructions, including acknowledging that the technology was still in its early stages and therefore still requires active oversight by HCPs to help in further implementation. Participants also emphasized the need for effective communication between users and Art developers to share experiences and to enable rapid tool adjustments when necessary.

#### Future Perspectives: Automatic Response System

The responses showed a wide range of opinions on the use of fully automatic response systems. Some participants considered it a promising idea, while others found it unacceptable. Several respondents indicated that such a system might only be appropriate under certain conditions, such as using it for simple questions only, adding a disclaimer that the message was generated by AI, improving the quality of the content, and increasing trust in the system among users. Some participants expressed concerns about the impersonal nature of automated messages and questioned who would be responsible for the content. Multiple people emphasized the importance of having HCPs review the messages before they are sent. Others mentioned the need to stay informed about what is being communicated to their patients, as they want to be up to date about relevant developments with their patients.

## Discussion

This study evaluated the effectiveness, feasibility, and barriers and facilitators to implement the Art GenAI tool. We found a shift from initial optimism surrounding Art use to more critical evaluations as users gained experience, reflected in both effectiveness and feasibility outcomes. With respect to effectiveness, early expectations were high, as indicated by the high scores on Clinical Efficiency outcomes in the PRE survey. However, mixed model analyses revealed growing dissatisfaction over time. Well-being outcomes showed no significant differences, and efficiency indicators showed that using Art did not appear to have impact on the time spent reviewing patient’s messages and drafting replies. In terms of feasibility, adoption of Art was low and declined over time across all departments, accompanied by decreases in user feedback and usability ratings. Analysis of barriers and facilitators of Art implementation revealed diverse user expectations and experiences. Identified themes mostly centered around the technology and adopters domains of the NASSS framework [31].

Facilitators included time savings, reduced mental demand, well-structured drafts, patient-friendly tones, and the ease of adjusting suggested replies. Interestingly, several barriers mirrored the facilitators, such as limited impact on time, increased mental demand, and style misalignment. Other barriers included incorrect or incomplete medical information and the need for verification by the users. Effective communication and education were considered essential to manage expectations and support implementation of Art in clinical practice. Other suggestions for improvement focused on offering customizable input, multiple draft messages, and a (personal) learning curve of Art. While some participants were open to an automatic response system if substantially improved, concerns about responsibility and preserving personal connection with patients were raised.

Previous work by Garcia et al. reported statistically significant reductions for task load and work exhaustion following implementation of Art [13]. Their study deployed a modified NASA Task Load Index, with four items instead of the original six. Similarly, Yadav and colleagues observed significant improvements in burnout, measured by the Mini Z 2.0 survey, and a significant reduction in NASA Task Load Index scores [15]. This contrasts with our findings, where Art showed limited influence on overall work exhaustion and task load. One possible explanation is that workload differences between settings may have influenced the perceived impact of Art.

For example, Garcia et al. documented 12,844 messages in five weeks for 162 participants, and Yadav et al. reported 1,700 messages over three months for 25 participants. In our study, the volume was 8,410 messages over six months for 237 participants, which is substantially lower per user than in U.S. contexts. While administrative workload is also a concern in Dutch healthcare [32], this may originate less from patient-provider messaging compared to other mentioned healthcare systems. Consequently, the potential for GenAI to reduce cognitive load may be less pronounced in our setting.

Adoption rates for Art have generally been low, averaging around 20% across studies. Reported adoption ranged from 12% [33] to 20% [13], with Yadav et al. and Mandal and colleagues documenting rates of 16.2% [15] and 19.4% [34] respectively. The only exception (at 58%) was Bootsma-Robroeks et al. [16], which was based on a clinician-only sample rather than our sample of physicians, nurses, and support staff. Our study observed an average of 16.7% across all users, starting at a higher level and declining over time.

Yadav et al. also reported initial high expectations regarding effectiveness (usefulness and cognitive burden) and feasibility (quality), followed by modest declines in the POST-survey [15]. These patterns mirror our findings and highlight the discrepancy between early optimism and actual experiences. In the future, we recommend incorporating user feedback systematically and early in the implementation process, for example by using iterative prompt engineering cycles [12]. Establishing a clear structure for how feedback is collected and acted upon can help address barriers and maintain user engagement and satisfaction over time [35–37]. As a concrete example for Art, integrating Text Assistant (Epic Systems Corporation, Verona, WI, USA) could directly address the style misalignment barrier and the need for customizable input. Text Assistant can rewrite sentences into a more formal, informal, or patient-friendly version, which would allow HCP to tailor the tone of their messages. This would operationalize user feedback directly and provide a practical step toward reducing style-related barriers.

Additionally, to support future implementation efforts, it will be essential to establish consensus on quality indicators and their thresholds that define when a GenAI tool provides meaningful added value within a specific clinical context. This could inform institutions to consider further development and implementation of GenAI tools in clinical practice and monitoring of the quality of GenAI tools’ output over time.

Qualitative data from Garcia et al. further revealed contrasting comments, with most concerns related to context accuracy and the varied impact on workflow efficiency [13]. Our findings reflect a similar pattern, as participants reported both facilitators, such as time savings and ease of adjusting drafts, and barriers, including the need for verification. These contrasting views may partly originate from differing initial expectations. Some users may have anticipated a complete, accurate draft ready for immediate use, while others expected a preliminary structure that could reduce cognitive load but still required medical review. Part of this discrepancy may also stem from differences between widely used web-based GenAI tools, which are more interactive, and Art, which is a static tool that does not learn from prior interactions. Other barriers and suggestions for improvement included concerns about tool reliability, lack of experience, and user enablement. These challenges are not unique to GenAI but reflect broader challenges in AI implementation. Farič et al. highlighted the clinician’s need to verify AI outputs and the risk of automation bias [39]. During interviews, radiologists stressed that the ultimate responsibility for clinical decisions should remain with them and not with AI-based clinical aids [39]. Swillens et al. reported comparable findings, noting that verification of AI-generated output was a barrier for implementing computational pathology algorithms [40]. To address these barriers, future implementation of GenAI should include clear communication of the intended purpose and functionality of GenAI tools, as well as explicit guidance on clinical responsibility. This is also in line with Article 4 of the EU AI Act [38], which emphasized the need for transparent communication and education regarding model capabilities and limitations during implementation.

### Strengths & Limitations

Using a hybrid type 1 effectiveness-implementation design was a strength of this study, as it allowed barriers and facilitators to be identified early in the implementation process [17]. This proactive approach provided valuable insights for future implementation of GenAI in healthcare. Another strength was the inclusion of a broad range of departments, both surgical and non-surgical, and users with different professions. This enhanced the diversity of perspectives and increased the relevance of findings across different clinical contexts. Although some participants had limited experience with the tool, their feedback still offered meaningful direction for successful implementation. Lastly, we used multiple time measurements, including POST-2, which added depth by capturing longer-term perceptions beyond initial impressions.

Several limitations should be acknowledged. First, the efficiency indicators included a different user population, consisting of both InBasket users with and without Art, which means that these findings should be interpreted with caution. Additionally, this study focused exclusively on HCPs’ perspective regarding usability. Future studies should also incorporate patient perspectives to understand how they perceive message generated with GenAI support.

Second, response rates varied substantially across the three surveys. The high response rate for the PRE survey can be explained by the initial requirement to complete the survey before gaining access to the tool. In contrast, POST-1 and POST-2 had lower participation, partly due to repetitive questions and limited motivation to complete follow-up surveys. For future studies, we recommend using a single POST measurement after 2-3 months to ensure participants have sufficient experience while reducing survey fatigue.

Third, the survey development followed a pragmatic approach. We deliberately opted for a concise and practical instrument rather than incorporating a very extensive survey instrument (e.g., the Unified Theory of Acceptance and Use of Technology, UTAUT [24]). Future evaluations could adopt a more robust methodology, as also emphasized by Hu et al. [12]. Standardized evaluation frameworks should be established through a more extensive literature review and consensus-building among professionals about the outcomes that should be included in the survey. For instance, outcomes surrounding bias and hallucinations in the safety and harm domain of the QUEST framework [41]. Additionally, self-efficacy and digital skills from the UTAUT could be included as an indicator of adoption and implementation [24].

Finally, fourth, validation results of the survey indicated that for some domains, factor loadings were low, and model fit was poor, raising concerns about the validity of the instrument. For example, the SUS scale showed a low CFI score, possibly because not all items were relevant to our tool. Although we included all items of the SUS due to its status as a validated instrument, future work should re-evaluate the use of existing instruments in the validation of GenAI. Also, poor factor loadings and model fit were observed at POST-2, likely due to the small sample size, which may have contributed to unreliable results at that time point. These findings suggest that certain constructs and instruments may require revision in future survey iterations and highlight the need for a standardized validation instrument for GenAI tools.

### Conclusions

Using a hybrid type 1 effectiveness-implementation design, this study presents an evaluation of a GenAI tool designed to support patient-provider communication in a non-English hospital. Findings revealed high initial expectations, but lower perceived effectiveness and feasibility in day-to-day clinical practices. This suggests that, in its current form, Art does not fully meet user expectations. Our results further indicated mixed perceptions on barriers and facilitators. Beyond technical accuracy and incorporating user feedback continuously, successful implementation will depend on clear communication about the tool’s intended purpose and limitations, along with guidance on clinical responsibilities. Future work should also aim to establish consensus-driven quality criteria and their thresholds that indicate when a GenAI tool achieves the level of performance needed to be integrated into routine clinical practice. These findings are critical for responsibly scaling GenAI applications in clinical workflows.

## Supporting information

Multimedia Appendix 1 - CHERRIES guidelines

Multimedia Appendix 2 - Supplementary figures and tables

## Acknowledgments

We would like to thank all respondents of the survey for their valuable contribution to this study. Additionally, we gratefully acknowledge the GenAI workgroup for their support in implementing the GenAI tool at Radboudumc. We thank Dani Reuvekamp and Roel Boumans for their contribution to the development of the survey. We also extend our appreciation to Anja van der Cruijsen from the Department of Dermatology for her input and assistance in recruiting respondents. Finally, we thank Prof. dr. Rosella Hermens and ir. Reinier Akkermans for sharing their expertise in implementation science and statistics, respectively.

## Data Availability

The datasets generated or analyzed during this study are not publicly available due to ethical and privacy considerations. Pseudonymized datasets are available from the corresponding author on reasonable request.

## Author’s Contributions

Conceptualization: TF (lead), JS (equal), AdV (supporting) Data curation: KB (lead), JS (supporting)

Formal analysis: KB (lead), AV (supporting), JS (supporting)

Funding acquisition: JS (lead), TF (equal), AdV (supporting)

Investigation: KB (lead), AV (supporting), JS (supporting)

Methodology: JS (lead), KB (supporting), AV (supporting), MAT (supporting), TF (supporting) Project administration: TF (lead), JS (equal)

Resources: TF (lead), JS (supporting), GvdB (supporting), CD (supporting), RD (supporting), BR (supporting), AS (supporting)

Software: RW (lead), TF (supporting)

Supervision: JS (lead), TF (equal), AV (supporting), AdV (supporting) Validation: KB (lead), AV (supporting), JS (supporting)

Visualization: KB (lead)

Writing – original draft: KB (lead), JS (supporting)

Writing – review & editing: KB (lead), AV (supporting), MAT (supporting), RW (supporting), GvdB (supporting), CD (supporting), RD (supporting), BR (supporting), AS (supporting), AdV (supporting), TF (supporting), JS (supporting).

## Abbreviations

GenAI: Generative Artificial Intelligence
HCP: Healthcare Professional
LLM: Large Language Model
EHR: Electronic Health Record
Art: Augmented Response Technology
ENT: Otorhinolaryngology or Ear, Nose and Throat
TAM: Technology Acceptance Model
SUS: System Usability Scale
NPS: Net Promotor Score
NASSS: Nonadoption, Abandonment, Scale-up, Spread, and Sustainability

