## Supplementary material for "AI-Generated Responses to Patient’s Messages: Effectiveness, Feasibility and Implementation": Multimedia Appendix 2 - Supplementary figures and tables

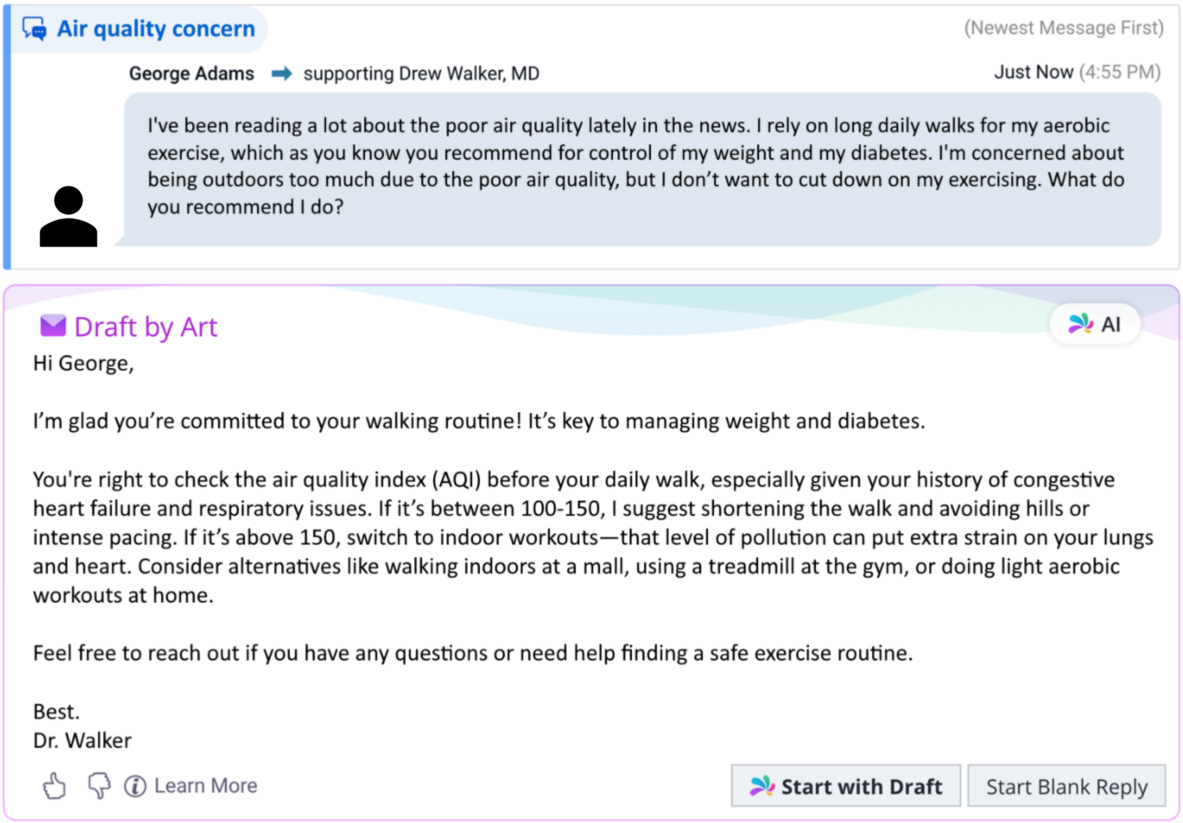


© 2026 Epic Systems Corporation

Figure S1. **Mock-up of the Art user interface.** All names and message content are fictional and used solely for demonstration purposes. Used with permission from Epic.

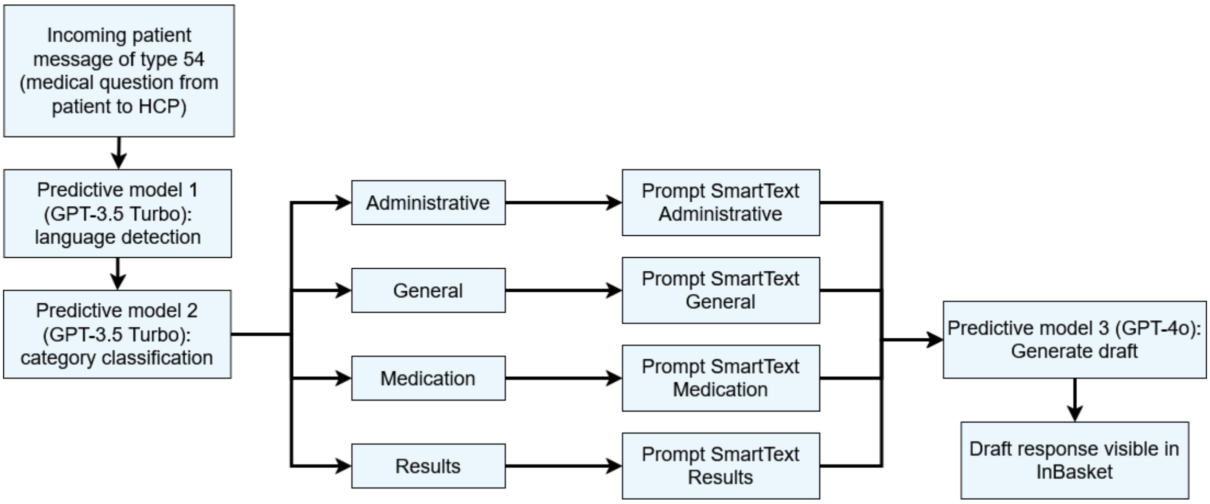


Figure S2. **Schematic of the technological workflow of Art.** Every incoming patient message of type 54 (medical question from patient to HCP) was classified into 4 categories using GPT-3.5 Turbo [1]: administrative, general, medication, and results. Each category was linked to a specific prompt. The draft message itself was automatically generated using the GPT-4o model [2] before being visible to users in InBasket.

Table S1. Overview and explanation of outcome measures collected from Epic.

| Measure | Metric | Meaning |
|---|---|---|
| **Efficiency indicators** | Review time | Time between reading a message and starting a reply. |
| | Draft time | Time spent composing a reply after initiating the draft (with either a blank reply or the generated draft). |
| | Messages with Art | Messages from users in the four included departments for which a draft was successfully generated by Art. |
| | Messages without Art | Messages either from departments not included in the study or from users in the four included departments where a draft could not be generated due to a technical error. |
| **Adoption indicators** | Number of generated drafts | How many draft replies were generated by Art. |
| | Number of drafts used | How many drafts generated by Art were used to start a reply with. This is regardless of how the content of the draft was edited by the user. |
| | Adoption rate | Number of drafts used divided by number of generated drafts. |
| | Number of positive feedback | Number of times a user clicked the positive feedback button. |
| | Number of constructive feedback | Number of times a user clicked the constructive feedback button, categorized into factual correctness, relevance, quality of patient guidance, incorrect name receiver, and length (too long or too short). |

Table S2. Structure of the survey, translated from Dutch.

| Domain | Scale | Question |
|---|---|---|
| **Characteristics** | Gender | What is your gender? |
| | Age | What is your age? |
| | Work experience (profession) | How many years of work experience do you have in your current profession? |
| | Work experience (total) | How many years of work experience do you have in total? (since obtaining your diploma) |
| | Experience with Epic | How long have you been working with EPIC? (in years) |
| | Experience with InBasket | How long have you been working with InBasket in Epic? (in years) |
| | Interest in GenAI within work environment | I am interested in GenAI within my work environment. (5-point Likert Scale: Strongly disagree – Strongly agree) |
| | Interest in GenAI outside work environment | I am interested in GenAI outside my work environment. (5-point Likert Scale: Strongly disagree – Strongly agree) |
| | Trust in AI [3–5] | What do you think about AI in general? (5-point Likert Scale: Strongly disagree – Strongly agree)<br>1)I am convinced that AI generally works well.;<br>2)The outputs of the AI-tools are predictable.;<br>3)I feel that I can count on AI.;<br>4)I feel safe that when I rely on AI, I will get correct/accurate information.;<br>5)I am wary of AI.;<br>6)I am suspicious about the intentions of AI.;<br>7)I have confidence in AI.;<br>8)I would follow the advice from AI. |
| | Department | Which department do you work in? |
| | Profession | What is your profession? |
| **Effectiveness: Well-being** | Work Exhaustion | Professional Fulfillment Index (PFI-WE) – work exhaustion scale [6] |
| | Task Load | NASA Task Load Index [7] |
| **Effectiveness: Clinical Efficiency** | Usefulness - TAM-PU [8] | To what extent do you agree with the following statements? (5-point Likert Scale: Strongly disagree – Strongly agree)<br>1)Using Art in practice helps me to accomplish tasks more quickly.;<br>2)Using Art in practice improves my work performance.;<br>3)Using Art in practice improves my productivity.;<br>4)Using Art in practice enhances my effectiveness in my job.;<br>5)Using Art in practice makes it easier to do my work.;<br>6)I find Art useful in my job. |
| | Effects [9] | Based on your experience with using Art, to what extent do you agree with the following |

| | | |
|---|---|---|
| | | statements? Art... (5-point Likert Scale: Strongly disagree – Strongly agree)<br>1)...is useful for answering patients' questions;<br>2)...saves time.<br>3)...improves the quality of answers to patient questions.<br>4)...improves the tone of answers to patient questions.<br>5)...ensures that I have to look up less information in Epic. (only for POST) |
| | Added value | What is the added value of using Art? List a maximum of 3 things. |
| **Feasibility: Use** | Art used | Have you used Art?<br>1) Yes<br>2) No |
| | If not Art used | What is the reason or what are the reasons why you did not use ART? List a maximum of 3 reasons. |
| | Weekly use (only POST) | How often have you used Art in the past week? (Each suggested draft counts as 1 time)<br>1) Not used; 2)1-5 times; 3)5-10 times; 4)10-15 times; 5)15-20 times; 6)>20 times |
| | Total use (only POST) | How often have you used Art in total? (Each suggested draft counts as 1 time.) |
| | Attitude towards Art (only PRE) – based on TAM-ATT [8] | To what extent do you agree with the following statements? (5-point Likert Scale: Strongly disagree – Strongly agree)<br>1) I think it's a good idea to start using Art.<br>2) I think it's a sensible idea to start using Art.<br>3) I like the idea of starting to use Art.<br>4) Using Art seems like a pleasant idea to me. |
| | Behavioral Intention to Use - TAM-BI [8] | I presently intend to use Art regularly in practice. (5-point Likert Scale: Strongly disagree – Strongly agree |
| **Feasibility: Usability of Art** | Ease of Use - TAM-PEOU [8] | To what extent do you agree with the following statements? (5-point Likert Scale: Strongly disagree – Strongly agree)<br>1)Learning to use Art has been easy for me.;<br>2)I find it easy to get Art to do what I want it to do.;<br>3)My interaction with Art is clear and understandable.;<br>4)I find Art to be flexible to interact with.;<br>5)It is easy for me to become skillful at using Art.;<br>6)I find Art easy to use. |
| | Output [10] | Based on your experience with using Art, to what extent do you agree with the following statements? Art messages... (5-point Likert Scale: Strongly disagree – Strongly agree)<br>1)...are accurate/correct.; |

| | | |
|---|---|---|
| | | 2)...do not contain any linguistic errors.;<br>3) ...are complete.;<br>4)...are reliable.;<br>5)...are good in terms of length. (only for POST) |
| | System Usability Scale (only POST) | System Usability Scale [11] |
| | Net Promotor Score (only POST) | Net Promotor Score [12] |
| **Implementation** | Barriers [13] | What bottlenecks are you currently experiencing when using Art? List a maximum of 3 bottlenecks. |
| | Facilitators [13] | What helps you to use Art effectively in practice now? List a maximum of 3 things. |
| | Suggestions for improvements | What could be improved about Art? List a maximum of 3 improvements. |
| **Future** | Future perspectives on Automatic Response System | Suppose that in the future, Art could be used to automatically respond to messages without you, as a healthcare professional, having to look at the suggested text. What would you think of this? |

Table S3. Cronbach's alpha coefficients for each scale across the three survey time points.

| Scale | PRE | POST-1 | POST-2 |
|---|---|---|---|
| **Work Exhaustion** | 0.78 | 0.80 | 0.52* |
| **Task Load** | 0.76 | 0.70 | 0.36* |
| **Usefulness** | 0.86 | 0.89 | 0.95 |
| **Effects** | 0.75 | 0.75 | 0.88 |
| **Attitude towards Art** | 0.93 | N/A | N/A |
| **Ease of Use** | 0.87 | 0.85 | 0.74 |
| **Output** | 0.80 | 0.75 | 0.78 |
| **System Usability Scale** | N/A | 0.74 | 0.79 |

*Cronbach's alpha coefficients below 0.7 generally indicate low internal consistency.

Table S4. Confirmative factor analysis results for each scale, including factor loading ranges, chi-square statistics ($\chi^2$), Comparative Fit Index (CFI), and Root Mean Square Error of Approximation (RMSEA).

| Scale | Survey | Loadings range | $\chi^2$ (p-value) | CFI | RMSEA |
|---|---|---|---|---|---|
| **Work Exhaustion** | PRE | 0.61 - 0.81 | 14.39 (p<.001) | 0.91 | 0.24* |
| | POST-1 | 0.60 - 0.91 | 8.41 (p=.01) | 0.91 | 0.27* |
| | POST-2 | –0.17 - 0.98 | 7.98 (p=.02) | 0.23* | 0.33* |
| **Task Load** | PRE | 0.20 - 0.79 | 4.09 (p=.91) | 1.02 | 0.0 |
| | POST-1 | 0.07 - 0.77 | 10.54 (p=.31) | 0.97 | 0.06 |
| | POST-2 | –0.30 - 0.85 | 7.88 (p=.55) | 1.14 | 0.0 |
| **Usefulness** | PRE | 0.57 - 0.86 | 14.71 (p=.10) | 0.98 | 0.08 |
| | POST-1 | 0.70 - 0.85 | 26.11 (p=.002) | 0.89* | 0.21* |
| | POST-2 | 0.82 - 0.93 | 28.91 (p<.001) | 0.89* | 0.28* |
| **Effects** | PRE | 0.60 - 0.71 | 16.31 (p<.001) | 0.88* | 0.26* |
| | POST-1 | 0.40 - 0.90 | 10.04 (p=0.07) | 0.92 | 0.15* |
| | POST-2 | 0.61 - 0.89 | 22.77 (p<.001) | 0.80* | 0.36* |
| **Attitude towards Art** | PRE | 0.85 - 0.90 | 1.08 (p=.58) | 1.00 | 0.0 |
| **Ease of Use** | PRE | 0.62 - 0.81 | 28.91 (p<.001) | 0.93 | 0.14* |
| | POST-1 | 0.45 - 0.89 | 14.84 (p=.10) | 0.95 | 0.12* |
| | POST-2 | 0.23 - 0.79 | 9.58 (p=.39) | 0.98 | 0.05 |
| **Output** | PRE | 0.56 - 0.94 | 3.14 (p=0.21) | 0.99 | 0.07 |
| | POST-1 | 0.29 - 0.81 | 3.34 (p=0.65) | 1.03 | 0.0 |
| | POST-2 | 0.39 - 0.80 | 8.43 (p=0.13) | 0.91 | 0.16* |
| **System Usability Scale** | POST-1 | 0.17 - 0.83 | 83.42 (p<.001) | 0.59* | 0.18* |
| | POST-2 | 0.21 - 0.83 | 80.74 (p<.001) | 0.58* | 0.22* |

* CFI values below 0.90 and RMSE values above 0.10 indicate poor model fit.

Table S5. Total utilization of Art per department during 6 months.

| | Total number of generated drafts | Total number of drafts used | Use (%) |
|---|---|---|---|
| **General** | 8,410 | 1,401 | 16.7% |
| **Dermatology** | 2,308 | 349 | 15.1% |
| **ENT** | 963 | 171 | 17.8% |
| **Pulmonology** | 2,210 | 598 | 27.1% |
| **Medical Oncology** | 2,559 | 273 | 10.7% |

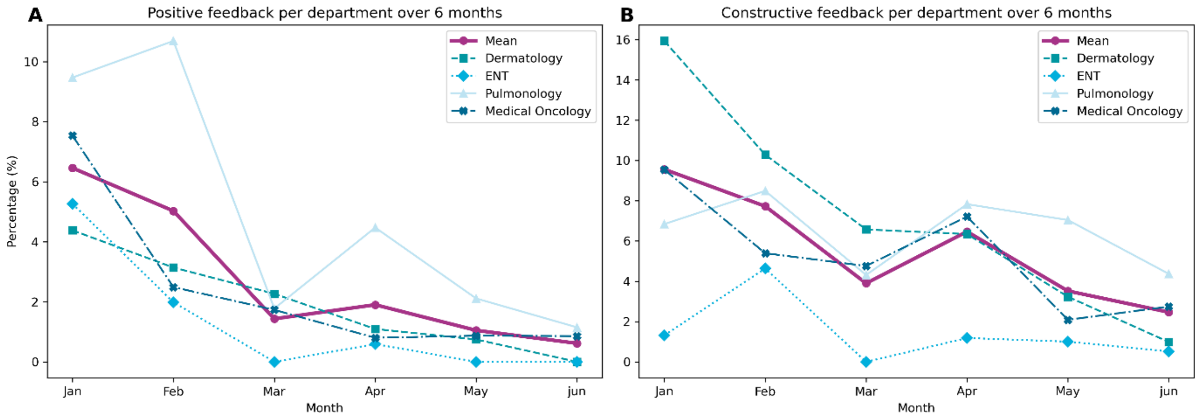


Figure S3. **Percentage of positive (A) and constructive (B) feedback reported per department over a six-month period.** The figures show the proportion of positive (A) and constructive (B) feedback relative to the total number of generated drafts per department, along with the overall average across all departments.

Table S6. Descriptive statistics for domain Well-being.

| | PRE | n | POST-1 | n | POST-2 | n |
|---|---|---|---|---|---|---|
| **Work Exhaustion** | 1.56 (0.63) | 108 | 1.60 (0.68) | 46 | 1.41 (0.37) | 29 |
| Department | | | | | | |
| *Dermatology* | 1.69 (0.75) | 31 | 1.53 (0.76) | 18 | 1.32 (0.28) | 7 |
| *ENT* | 1.68 (0.52) | 15 | 1.71 (0.43) | 6 | 1.42 (0.56) | 6 |
| *Pulmonology* | 1.37 (0.46) | 19 | 1.77 (0.61) | 11 | 1.45 (0.29) | 11 |
| *Medical Oncology* | 1.50 (0.63) | 43 | 1.50 (0.75) | 11 | 1.45 (0.48) | 5 |
| Profession | | | | | | |
| *Physician* | 1.47 (0.63) | 56 | 1.64 (0.70) | 23 | 1.45 (0.35) | 20 |
| *Nurse* | 1.63 (0.66) | 40 | 1.69 (0.69) | 16 | 1.33 (0.43) | 9 |
| *Support staff* | 1.71 (0.57) | 12 | 1.29 (0.57) | 7 | | 0 |
| **Task Load** | 33.93 (17.78) | 108 | 38.06 (15.52) | 46 | 35.14 (10.80) | 29 |
| Department | | | | | | |
| *Dermatology* | 39.70 (17.96) | 31 | 39.03 (14.13) | 18 | 39.05 (12.89) | 7 |
| *ENT* | 36.72 (16.70) | 15 | 36.94 (8.11) | 6 | 30.42 (10.98) | 6 |
| *Pulmonology* | 33.99 (18.96) | 19 | 42.88 (10.29) | 11 | 34.55 (9.82) | 11 |
| *Medical Oncology* | 28.76 (16.53) | 43 | 32.27 (23.25) | 11 | 36.67 (10.59) | 5 |
| Profession | | | | | | |
| *Physician* | 37.17 (16.60) | 56 | 42.50 (14.41) | 23 | 35.79 (10.70) | 20 |
| *Nurse* | 30.23 (18.52) | 40 | 32.40 (17.13) | 16 | 33.70 (11.55) | 9 |
| *Support staff* | 31.11 (19.20) | 12 | 36.43 (12.11) | 7 | | 0 |

Table S7. Descriptive statistics for domain Clinical Efficiency.

| | PRE | n | POST-1 | n | POST-2 | n |
|---|---|---|---|---|---|---|
| **Usefulness** | 3.71 (0.53) | 108 | 3.37 (0.66) | 46 | 3.13 (0.89) | 29 |
| Department | | | | | | |
| *Dermatology* | 3.80 (0.76) | 31 | 3.34 (0.58) | 18 | 3.55 (0.34) | 7 |
| *ENT* | 3.86 (0.32) | 15 | 3.67 (0.80) | 6 | 3.31 (0.55) | 6 |
| *Pulmonology* | 3.69 (0.40) | 19 | 3.27 (0.72) | 11 | 3.00 (1.20) | 11 |
| *Medical Oncology* | 3.60 (0.42) | 43 | 3.33 (0.70) | 11 | 2.60 (0.77) | 5 |
| Profession | | | | | | |
| *Physician* | 3.76 (0.61) | 56 | 3.36 (0.72) | 23 | 3.08 (0.86) | 20 |
| *Nurse* | 3.65 (0.42) | 40 | 3.39 (0.49) | 16 | 3.22 (0.99) | 9 |
| *Support staff* | 3.64 (0.44) | 12 | 3.36 (0.85) | 7 | | 0 |
| **Effects** | 3.58 (0.62) | 108 | 3.33 (0.60) | 46 | 3.17 (0.93) | 29 |
| Department | | | | | | |
| *Dermatology* | 3.70 (0.74) | 31 | 3.47 (0.39) | 18 | 3.54 (0.28) | 7 |
| *ENT* | 3.80 (0.41) | 15 | 3.43 (0.82) | 6 | 3.67 (0.99) | 6 |
| *Pulmonology* | 3.55 (0.63) | 19 | 3.16 (0.67) | 11 | 3.02 (1.12) | 11 |
| *Medical Oncology* | 3.44 (0.55) | 43 | 3.20 (0.67) | 11 | 2.40 (0.37) | 5 |
| Profession | | | | | | |
| *Physician* | 3.61 (0.72) | 56 | 3.17 (0.68) | 23 | 3.07 (0.90) | 20 |
| *Nurse* | 3.54 (0.49) | 40 | 3.44 (0.50) | 16 | 3.40 (1.00) | 9 |
| *Support staff* | 3.60 (0.49) | 12 | 3.57 (0.42) | 7 | | 0 |

Table S8. Descriptive statistics for domain Use of Art.

| | PRE | n | POST-1 | n | POST-2 | n |
|---|---|---|---|---|---|---|
| **Art used at least once*** | | | | | | |
| Yes | N/A | | 46 (79.3%) | | 29 (78.4%) | |
| No | N/A | | 12 (20.7%) | | 8 (21.6%) | |
| **How often used in the past week?*** | | | | | | |
| 0 times | N/A | | 5 (8.6%) | | 0 | |
| 1-5 times | N/A | | 36 (62.1%) | | 21 (56.8%) | |
| 5-10 times | N/A | | 2 (3.4%) | | 5 (13.5%) | |
| 10-15 times | N/A | | 1 (1.7%) | | 2 (5.4%) | |
| 15-20 times | N/A | | 0 | | 0 | |
| >20 times | N/A | | 2 (3.4%) | | 1 (2.7%) | |
| **Total use** | N/A | | 9.65 (12.26) | 46 | 16.07 (21.72) | 29 |
| **Attitude towards Art** | 3.84 (0.64) | 107 | N/A | | N/A | |
| Department | | | | | | |
| *Dermatology* | 4.02 (0.71) | 31 | N/A | | N/A | |
| *ENT* | 3.95 (0.34) | 15 | N/A | | N/A | |
| *Pulmonology* | 3.81 (0.51) | 18 | N/A | | N/A | |
| *Medical Oncology* | 3.69 (0.69) | 43 | N/A | | N/A | |
| Profession | | | | | | |
| *Physician* | 3.98 (0.60) | 56 | N/A | | N/A | |
| *Nurse* | 3.62 (0.66) | 39 | N/A | | N/A | |
| *Support* | 3.94 (0.54) | 12 | N/A | | N/A | |
| **Behavioral Intention to Use⁺** | 4.0 (0.00) | 108 | 4.0 (0.00) | 46 | 4.0 (1.00) | 29 |
| Department | | | | | | |
| *Dermatology* | 4.0 (0.00) | 31 | 4.0 (0.00) | 18 | 4.0 (0.00) | 7 |
| *ENT* | 4.0 (0.50) | 15 | 4.0 (0.75) | 6 | 4.0 (0.00) | 6 |
| *Pulmonology* | 4.0 (0.00) | 19 | 4.0 (0.00) | 11 | 4.0 (1.00) | 11 |
| *Medical Oncology* | 4.0 (0.50) | 43 | 4.0 (0.00) | 11 | 3.0 (2.00) | 5 |
| Profession | | | | | | |
| *Physician* | 4.0 (0.25) | 56 | 4.0 (0.00) | 23 | 4.0 (1.00) | 20 |
| *Nurse* | 4.0 (1.00) | 40 | 4.0 (0.00) | 16 | 4.0 (0.00) | 9 |
| *Support staff* | 4.0 (1.00) | 12 | 4.0 (0.00) | 7 | | 0 |

* Number of times and percentage;

⁺ Behavioral Intention to Use consisted of only one item in the scale. Therefore, the median and interquartile range (IQR) are reported instead of the mean and standard deviation (SD).

Table S9. Descriptive Statistics for domain Usability of Art.

| | PRE | n | POST-1 | n | POST-2 | n |
|---|---|---|---|---|---|---|
| **Ease of Use** | 3.58 (0.53) | 108 | 3.59 (0.58) | 46 | 3.34 (0.57) | 29 |
| Department | | | | | | |
| *Dermatology* | 3.69 (0.48) | 31 | 3.66 (0.59) | 18 | 3.43 (0.57) | 7 |
| *ENT* | 3.69 (0.63) | 15 | 3.61 (0.56) | 6 | 3.53 (0.29) | 6 |
| *Pulmonology* | 3.49 (0.42) | 19 | 3.50 (0.59) | 11 | 3.38 (0.74) | 11 |
| *Medical Oncology* | 3.51 (0.57) | 43 | 3.58 (0.62) | 11 | 2.93 (0.25) | 5 |
| Profession | | | | | | |
| *Physician* | 3.64 (0.54) | 56 | 3.63 (0.58) | 23 | 3.32 (0.55) | 20 |
| *Nurse* | 3.45 (0.47) | 40 | 3.49 (0.54) | 16 | 3.41 (0.66) | 9 |
| *Support staff* | 3.76 (0.62) | 12 | 3.71 (0.69) | 7 | | 0 |
| **Output** | 3.40 (0.50) | 108 | 3.21 (0.51) | 46 | 3.21 (0.58) | 29 |
| Department | | | | | | |
| *Dermatology* | 3.48 (0.58) | 31 | 3.19 (0.50) | 18 | 3.37 (0.35) | 7 |
| *ENT* | 3.40 (0.41) | 15 | 3.30 (0.30) | 6 | 3.50 (0.30) | 6 |
| *Pulmonology* | 3.34 (0.53) | 19 | 3.04 (0.60) | 11 | 2.98 (0.82) | 11 |
| *Medical Oncology* | 3.38 (0.46) | 43 | 3.36 (0.54) | 11 | 3.16 (0.26) | 5 |
| Profession | | | | | | |
| *Physician* | 3.42 (0.55) | 56 | 3.09 (0.59) | 23 | 3.12 (0.58) | 20 |
| *Nurse* | 3.39 (0.43) | 40 | 3.31 (0.39) | 16 | 3.42 (0.56) | 9 |
| *Support staff* | 3.38 (0.52) | 12 | 3.37 (0.42) | 7 | | 0 |
| **System Usability Scale (SUS)** | N/A | | 67.01 (9.00) | 46 | 65.52 (11.01) | 29 |
| Department | | | | | | |
| *Dermatology* | N/A | | 69.03 (6.81) | 18 | 63.93 (8.76) | 7 |
| *ENT* | N/A | | 64.17 (9.44) | 6 | 70.42 (6.97) | 6 |
| *Pulmonology* | N/A | | 65.68 (10.55) | 11 | 59.50 (11.37) | 11 |
| *Medical Oncology* | N/A | | 66.59 (10.74) | 11 | 66.59 (13.48) | 5 |
| Profession | | | | | | |
| *Physician* | N/A | | 68.91 (8.59) | 23 | 64.75 (9.66) | 20 |
| *Nurse* | N/A | | 63.44 (9.99) | 16 | 67.22 (14.06) | 9 |
| *Support staff* | N/A | | 68.93 (5.93) | 7 | | 0 |
| **Net Promotor Score (NPS)** | N/A | | –13.04 | 46 | –37.93 | 29 |
| Department | | | | | | |
| *Dermatology* | N/A | | –5.56 | 18 | 0.00 | 7 |
| *ENT* | N/A | | –33.33 | 6 | –16.67 | 6 |
| *Pulmonology* | N/A | | –18.18 | 11 | –80.00 | 11 |
| *Medical Oncology* | N/A | | –9.09 | 11 | –54.55 | 5 |
| Profession | | | | | | |
| *Physician* | N/A | | –26.09 | 23 | –45.00 | 20 |
| *Nurse* | N/A | | –6.25 | 16 | –22.22 | 9 |
| *Support staff* | N/A | | 14.29 | 7 | | 0 |

Table S10. Changes over time across all survey scales.

| Scale | Time interval | Effect | 95% CI* | p-value | Cohen's d* |
|---|---|---|---|---|---|
| **Work Exhaustion** | PRE vs POST-1 | 0.04 | –0.11 to 0.19 | p=.62 | 0.06 (small) |
| | PRE vs POST-2 | –0.13 | –0.32 to 0.06 | p=.17 | –0.21 (small) |
| **Task Load** | PRE vs POST-1 | 3.00 | –1.36 to 7.36 | p=.18 | 0.17 (small) |
| | PRE vs POST-2 | 0.96 | –4.52 to 6.43 | p=.73 | 0.05 (small) |
| **Usefulness** | PRE vs POST-1 | –0.39 | –0.58 to –0.20 | p<.001 | –0.73 (medium to large) |
| | PRE vs POST-2 | –0.57 | –0.80 to –0.34 | p<.001 | –1.07 (large) |
| **Effects** | PRE vs POST-1 | –0.33 | –0.51 to 0.15 | p<.001 | –0.54 (medium) |
| | PRE vs POST-2 | –0.37 | –0.60 to –0.15 | p<.001 | –0.61 (medium to large) |
| **Intention to Use** | PRE vs POST-1 | –0.22 | –0.44 to 0.01 | p=.06 | –0.31 (small to moderate) |
| | PRE vs POST-2 | –0.30 | –0.58 to –0.03 | p=.03 | –0.44 (moderate) |
| **Ease of Use** | PRE vs POST-1 | –0.02 | –0.19 to 0.14 | p=.77 | –0.05 (small) |
| | PRE vs POST-2 | –0.23 | –0.43 to –0.03 | p=.03 | –0.43 (moderate) |
| **Output** | PRE vs POST-1 | –0.22 | –0.38 to –0.07 | p=.005 | –0.44 (moderate) |
| | PRE vs POST-2 | –0.17 | –0.36 to 0.02 | p=.09 | –0.34 (small to moderate) |
| **System Usability Scale** | POST-1 vs POST-2 | –1.49 | –6.26 to 3.27 | p=.54 | –0.17 (small) |

* CI = Confidence Interval; Cohen's d = Effect size.
